# Validation of home oxygen saturations as a marker of clinical deterioration in patients with suspected COVID-19

**DOI:** 10.1101/2020.11.06.20225938

**Authors:** Matthew Inada-Kim, Francis P. Chmiel, Michael J. Boniface, Helen Pocock, John J. M. Black, Charles D. Deakin

## Abstract

**Background:** The early identification of deterioration in suspected COVID-19 patients managed at home enables a more timely clinical intervention, which is likely to translate into improved outcomes. We undertook an analysis of COVID-19 patients conveyed by ambulance to hospital to investigate how oxygen saturation and measurements of other vital signs correlate to patient outcomes, to ascertain if clinical deterioration can be predicted with simple community physiological monitoring.

**Methods:** A retrospective analysis of routinely collected clinical data relating to patients conveyed to hospital by ambulance was undertaken. We used descriptive statistics and predictive analytics to investigate how vital signs, measured at home by ambulance staff from the South Central Ambulance Service, correlate to patient outcomes. Information on patient comorbidities was obtained by linking the recorded vital sign measurements to the patient’s electronic health record at the Hampshire Hospitals NHS Foundation Trust. ROC analysis was performed using cross-validation to evaluate, in a retrospective fashion, the efficacy of different variables in predicting patient outcomes.

**Results:** We identified 1,080 adults with a COVID-19 diagnosis who were conveyed by ambulance to either Basingstoke & North Hampshire Hospital or the Royal Hampshire County Hospital (Winchester) between March 1^st^ and July 31^st^ and whose diagnosis was clinically confirmed at hospital discharge. Vital signs measured by ambulance staff at first point of contact in the community correlated with patient short-term mortality or ICU admission. Oxygen saturations were the most predictive of mortality or ICU admission (AUROC 0.772 (95 % CI: 0.712-0.833)), followed by the NEWS2 score (AUROC 0.715 (95 % CI: 0.670-0.760), patient age (AUROC 0.690 (95 % CI: 0.642-0.737)), and respiration rate (AUROC 0.662 (95 % CI: 0.599-0.729)). Combining age with the NEWS2 score (AUROC 0.771 (95 % CI: 0.718-0.824)) or the measured oxygen saturation (AUROC 0.820 (95 % CI: 0.785-0.854)) increased the predictive ability but did not reach significance.

**Conclusions:** Initial oxygen saturation measurements (on air) for confirmed COVID-19 patients conveyed by ambulance correlated with short-term (30-day) patient mortality or ICU admission, AUROC: 0.772 (95% CI: 0.712-0.833). We found that even small deflections in oxygen saturations of 1-2% below 96% confer an increased mortality risk in those with confirmed COVID at their initial community assessments.

## INTRODUCTION

COVID-19 presents the biggest global healthcare challenge of our generation. As of October 2020, COVID-19 associated mortality stands at 43,205 in the UK and, after a lull over the lockdown period, infection rates in the UK are on an upward trajectory again.[1] COVID-19 presents a number of challenges in identifying optimal management pathways, not only in terms of the clinical care itself, but also identifying the stage at which hospital admission is necessary. Traditional management pathways involving paramedic assessment and ambulance conveyance to the Emergency Department for further review have proven impractical, not only because of the large numbers of patients involved, but because of the need to minimise contact of COVID-19 patients with others. Most patients who become symptomatic do so in a home environment where the majority will remain. In terms of optimising outcome, there is a need to understand what symptoms and signs in this environment are prognostic indicators of potential deterioration. The national recommendation for the implementation of COVID virtual wards recently announced by NHS England,[2] ushers in a novel approach of empowering patients through giving symptomatic, at risk patients a pulse oximeter and a toolkit for self-monitoring at home. It is hoped that this will enable the earlier recognition of deterioration in COVID-19 patients and potentially improved outcomes.

In most cases of bacteria and non-COVID pneumonia, breathlessness appears relatively early in the disease and ahead of any significant hypoxia. The challenge with assessing COVID-19 severity is that asymptomatic hypoxia often precedes breathlessness and by the time symptoms of breathlessness occur, hypoxia may be significant.[3] The ability to detect this asymptomatic hypoxia before patients experience shortness of breath is critical for preventing the pneumonia progressing to a life-threatening state. The key is to be able to detect this initial drop in oxygen saturation levels so that patients infected with COVID-19 who begin to suffer from pneumonia in the community can be detected early and conveyed to hospital for further treatment, reducing the risk of mechanical ventilation with its associated high mortality.[4] Although some studies have reported the relationship between oxygen saturation and outcome on presentation to the Emergency Department, we are not aware of any studies that have reported the relationship between home oxygen saturations and outcome. Patients who on assessment are severely hypoxic are clearly in need of ambulance conveyance and hospital treatment, but by far the majority of patients with Covid-like symptoms seen and assessed by the ambulance service have relatively normal or near-normal oxygen saturations. These patients have generally not been conveyed and have been managed at home, but it has become apparent that even relatively minor derangements in oxygen saturations may be an early warning indicator for disease progression and the subsequent need for critical care. Use of oxygen saturation as an indicator of disease severity may therefore underestimate the risk of leaving patients at home after ambulance assessment. National case fatality rates (CFR) (ratio of deaths to total cases) have shown a strong inverse correlation between target oxygen saturation levels of 90-98% [5] suggesting that leaving even mild derangements in oxygen saturation untreated can be detrimental to outcome.

Understanding the prognostic implications of oxygen saturation when first measured by ambulance paramedics on patients at home would enable safe and effective triage and potentially improve outcome through early identification of those most at risk of disease progression. Two small studies have suggested the utility of home oxygen monitoring for COVID-19 patients discharged from hospital,[6, 7] but no studies to our knowledge have used home oxygen saturation as a trigger for initial hospital assessment. With second waves of COVID-19 sweeping most European countries, there is an urgent need to establish the prognostic significance of initial oxygen saturation to enable effective triage and optimise the use of limited healthcare resources, not only for those with COVID-19, but for the far greater majority with non-COVID-19 illness who have been deprived of timely healthcare as a consequence.

We therefore undertook a retrospective review of patients accessing a major UK ambulance service with symptoms suggestive of COVID-19 who were conveyed to hospital and correlated their initial oxygen saturations measured at home with their eventual outcome. These were compared with the standard NEWS2 patient score, as used by all UK ambulance services to identify the deteriorating patient.[8]

## METHODS

### Study Design

We conducted a retrospective cohort analysis of adult patients (aged 18 years of older) initially assessed and conveyed by personnel from South Central Ambulance Service (SCAS) to the Emergency Department at one of the two hospitals within north Hampshire; Basingstoke & North Hampshire Hospital, or the Royal Hampshire County Hospital (Winchester).

All calls to the ambulance service, both emergency (999) and urgent (111) are triaged using NHS Pathways telephone script (release 19). We analysed ambulance conveyances occurring between 1^st^ March to 31^st^ July 2020,to determine suspect COVID-19 among ambulance conveyances at initial time of contact by the call taker or ambulance crew, each patient record was reviewed for inclusion of at least one of the following four identifiers:

1. Those in who the ambulance call taker had classified the call as ‘COVID– Respiratory Distress’
2. Those where the Patient Clinical Record (PCR) listed the ‘Presenting complaint’ as ‘Suspected COVID-19’.
3. Those where the PCR free text for the ‘Presenting complaint’ contained the word ‘COVID’
4. Those where the PCR narrative in the free text field summarising the symptoms and their details completed by the paramedic contained the word ‘COVID’.

Conveyances from these suspect COVID-19 patients were then linked to their subsequent hospital attendance. Confirmed COVID-19 cases were then identified from these suspect cases by selecting those only those with a confirmed COVID-19 diagnosis in their discharge summary (i.e., the presence of a U07.1 or U07.2 ICD10 code).

All patients in known palliative care pathways were excluded from data analysis because their care did not follow standard care pathways.

### Study setting

SCAS is a provider of emergency 999 care in the counties of Hampshire, Berkshire, Buckinghamshire and Oxfordshire and covers a total of 3554 sq. miles (9205 km^2^). The service receives approximately 500,000 emergency and urgent calls annually. SCAS covers a residential population of approximately 4.0 million inhabitants in a mix of urban and rural areas. The north Hampshire region forms part of the area covered by SCAS and comprises a residential population of approximately 306,000.[9]

### Data collection

The initial oxygen saturation reading (SpO2) on air recorded by the attending ambulance crew (prior to any exercise or step test) and the NEWS2 score of patients fulfilling the inclusion criteria were collected from the ambulance service PCR. (NEWS2 score is calculated using the following seven variables: systolic blood pressure, heart rate, respiratory rate, temperature, oxygen saturation, supplemental oxygen administration, and level of consciousness - https://www.england.nhs.uk/ourwork/clinical-policy/sepsis/nationalearlywarningscore.)

Patient outcome was obtained by linking the ambulance and hospital clinical records by NHS number. Primary endpoint was death at 30 days. Secondary endpoints were hospital ICU admission and/or death at 24hours, 48 hours, 5 days and 30 days.

### Data analysis

Analysis was performed in Python, primarily making use of the statsmodels library. Confidence intervals on observed mortality rates were estimated using the Wilson score interval. Where relevant, significance of the difference between two observed mortality rates was tested using a two-population proportions z-test with the null hypothesis that the two-population proportions are equal.

To evaluate how predictive individual variables (e.g., oxygen saturation) and combinations of variables (e.g., oxygen saturation with age) were of 30-day mortality, we performed Receiving Operator Characteristics curve analysis (Table 2 and Table 3). In the univariate analysis, we assume a patient’s mortality risk is a linear function of the respective variable (where negative or positive correlation with mortality is assessed by clinical judgement) and calculated the ROC curve corresponding to if this variable alone was used to predict a patients mortality risk. When using a predictor which used age and either NEWS2 or oxygen saturation (bottom two rows in Table 3), the predicted risk for a patient is equal to the mean mortality rate for patients in the training data (discussed in the following) with similar age and NEWS2 / oxygen saturation as the respective patient, the bins used to define similarity are displayed in Supplementary Tables 1 and 2. We present either the sensitivity and specificity or the Area Under the Receiving Operator Characteristic curve (AUROC). The AUROC provides an estimate of the degree to which the predictor can discern between whether a patient dies within 30 days of conveyance or not, it can take values between 0.5 and 1.0. An AUROC of 0.5 corresponds to randomly guessing which patient will die within 30 days and an AUROC of 1.0 corresponds to a perfect classifier - it can predict, without error, who will die within 30-days of conveyance. Confidence intervals are estimated using a 10-fold cross validation procedure, we calculated the AUROC on the 10 (mutually exclusive) subsets of the data (the ‘validation’ data) and then report the mean AUROC alongside the standard error (at 95 % confidence) of the mean AUROC for these 10-folds. When either missing values need to be mean imputed (asterisks in Table 3) or values ‘learned’ from the data (i.e., for the bottom two models in Table 3), these values are calculated using the remaining 90 % of the data (the ‘training’ data) for each validation fold to avoid any leakage between our training and validation data.

## RESULTS

A total of 19,868 patients were assessed at home and subsequently conveyed by ambulance to North Hampshire Hospitals during the study period. The call handler or ambulance crew identified 2,257 suspect COVID-19 cases and of these we identified 1,209 adults as having a confirmed diagnosis of COVID-19 (U07.1 or U07.2 coded in the patients discharge summary). Those under palliative care (112 patients) and those with no initial oxygen saturation measurement on air recorded (17 patients) were excluded. In total, data was available from 1,080 patient records and of these, the complete records of vital signs were recorded at home by paramedics for 896 patients (Table 1).

**Table 1:** Number of vital sign measurements missing and the number of complete records from 1,080 patient records. ACVPU = <u>a</u>lert, <u>c</u>onfused, responding to <u>v</u>oice, responding to <u>p</u>ain, <u>u</u>nresponsive. Oxygen saturations were not missing for any patients as those with missing values had been excluded (n=17). Overall, records were complete for 83% of cases.

| Vital sign | Number missing | Percent missing |
| --- | --- | --- |
| Heart rate | 10 | 0.9 |
| Systolic blood pressure | 100 | 9.3 |
| Respiration rate | 120 | 11.1 |
| Oxygen Saturation (on air) | 0 | 0 |
| Temperature | 150 | 13.9 |
| ACVPU | 125 | 11.6 |
| Complete Records | 896 | 83.0 |

Age and known co-morbidities of, diabetes, chronic obstructive pulmonary disease (COPD), and cancer were examined in relation to mortality. While age displayed a clear correlation to 30-day mortality (Figure 1), patients either under cancer care (n=31) or with diabetes (n=235) were not found to be at significantly higher risk of 30-day mortality within the statistical power of this dataset. Although the observed 30-day mortality rate was higher for patients with COPD (mortality rate 14.1 %) compared to patients without COPD (mortality rate of 6.9 %), statistical significance was not reached (p=0.062).

**Figure 1:**
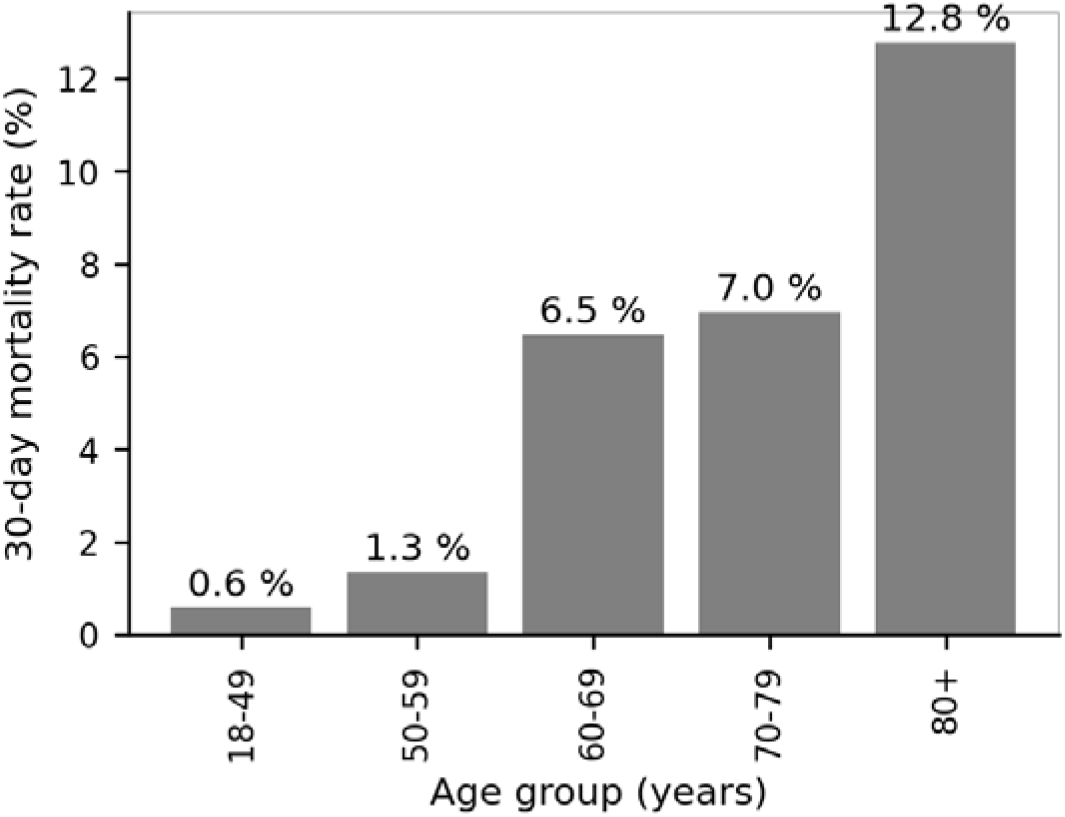
Observed 30-day mortality rate by age group for suspected COVID-19 patients conveyed by ambulance. Annotations (above bars) display the observed 30-day mortality rate for the respective group.

Oxygen saturation was found to correlate with mortality rate for both short- and longer-term outcomes (Figure 2 a), with lower initial oxygen saturation readings being associated with a higher mortality rate. In Figure 2 b we display the correlation between the observed 30-day mortality rate and initial oxygen saturation in detail, which evidently displays a non-linear relationship. In Table 2 we display the breakdown of our retrospective ROC analysis for using oxygen saturation as a binary triage tool (i.e., hospitalize or not) for different cut-offs (rows in Table 2). While the sensitivity vs specificity trade-off needs to be determined by the clinical context, this demonstrates that oxygen saturation is moderately discriminative for several cut-offs. For example, for a cut-off of 94 % or below, the sensitivity is 0.713 (95 % CI: 0.686-0.739) and the specificity is 0.723 (95 % CI: 0.705-0.742). Finally, we present comparisons of the results of ROC analysis for different variables measured in the community by paramedics (Table 3).

**Table 2:**
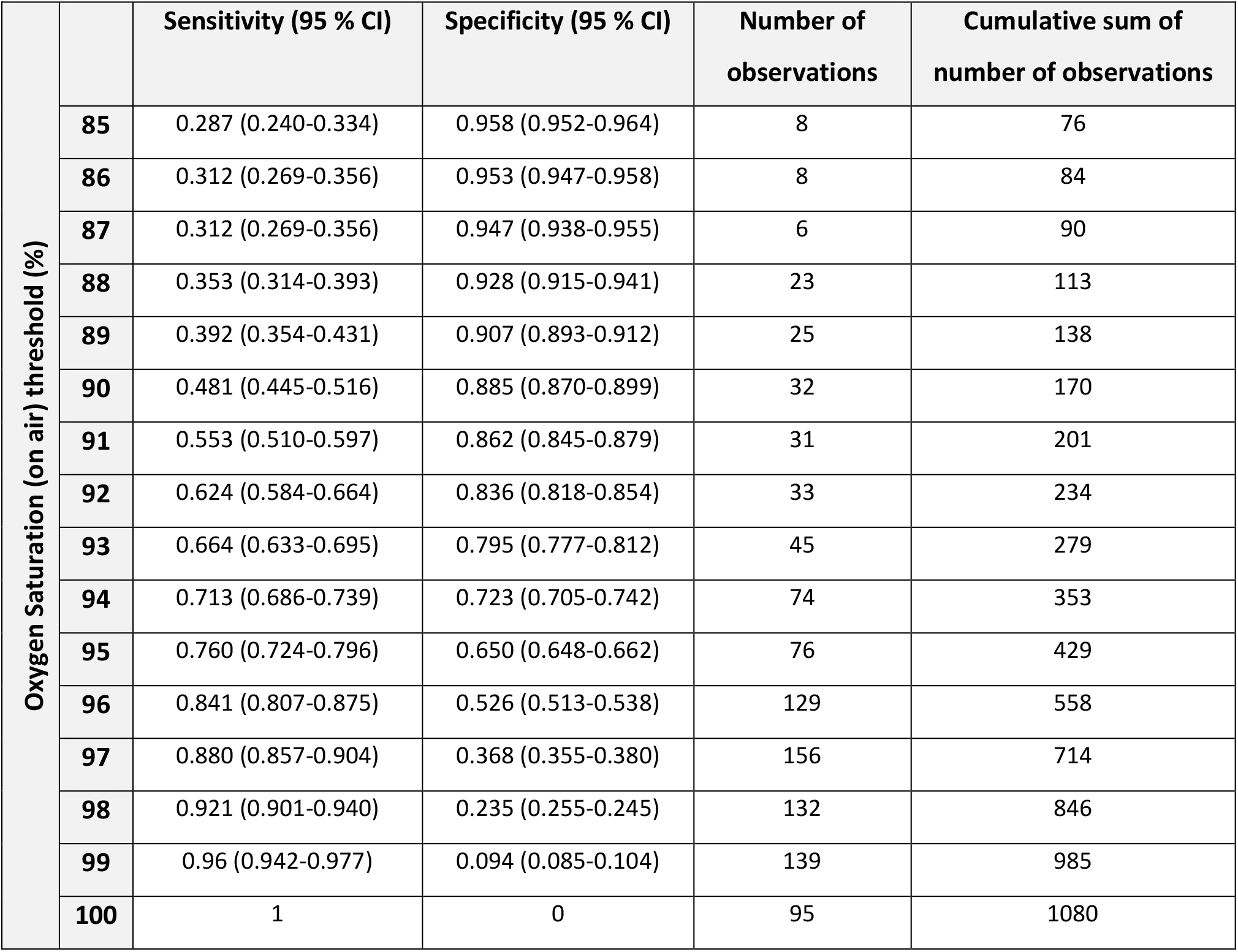
Evaluation of initial oxygen saturation measured by paramedics in COVID-19 patients in the community used as a binary classifier for predicting 30-day mortality or ICU admission. Each row denotes a different threshold for determining those at risk of death. We display the sensitivity and specificity for each threshold, equivalent to all possible intersections of the receiving operator curve using thresholds between 85 % and 100 %. In total 68 patients had an oxygen saturation of 84 % or less (not shown). The column on the far right denotes the cumulative sum of the number of observations of the given oxygen saturation (row) or below. For example, 76 patients had an oxygen saturation of 85 % or less recorded (top row) and 429 patients had an oxygen saturation of 95 % or less recorded.

| Oxygen Saturation (on air) threshold (%) |  | Sensitivity (95 % CI) | Specificity (95 % CI) | Number of observations | Cumulative sum of number of observations |
| --- | --- | --- | --- | --- | --- |
|  | 85 | 0.287 (0.240-0.334) | 0.958 (0.952-0.964) | 8 | 76 |
|  | 86 | 0.312 (0.269-0.356) | 0.953 (0.947-0.958) | 8 | 84 |
|  | 87 | 0.312 (0.269-0.356) | 0.947 (0.938-0.955) | 6 | 90 |
|  | 88 | 0.353 (0.314-0.393) | 0.928 (0.915-0.941) | 23 | 113 |
|  | 89 | 0.392 (0.354-0.431) | 0.907 (0.893-0.912) | 25 | 138 |
|  | 90 | 0.481 (0.445-0.516) | 0.885 (0.870-0.899) | 32 | 170 |
|  | 91 | 0.553 (0.510-0.597) | 0.862 (0.845-0.879) | 31 | 201 |
|  | 92 | 0.624 (0.584-0.664) | 0.836 (0.818-0.854) | 33 | 234 |
|  | 93 | 0.664 (0.633-0.695) | 0.795 (0.777-0.812) | 45 | 279 |
|  | 94 | 0.713 (0.686-0.739) | 0.723 (0.705-0.742) | 74 | 353 |
|  | 95 | 0.760 (0.724-0.796) | 0.650 (0.648-0.662) | 76 | 429 |
|  | 96 | 0.841 (0.807-0.875) | 0.526 (0.513-0.538) | 129 | 558 |
|  | 97 | 0.880 (0.857-0.904) | 0.368 (0.355-0.380) | 156 | 714 |
|  | 98 | 0.921 (0.901-0.940) | 0.235 (0.255-0.245) | 132 | 846 |
|  | 99 | 0.96 (0.942-0.977) | 0.094 (0.085-0.104) | 139 | 985 |
|  | 100 | 1 | 0 | 95 | 1080 |

**Table 3:** Area Under Receiver Operator Curves (AUROC) calculated for isolated physiological variables, the composite NEWS2 score, and composite models including either NEWS2 or Oxygen saturation and age with mortality at 30-days. The mean validation AUROC across the validation folds is presented and 95 % confidence intervals are estimated by the standard error of the AUROCs across the 10-folds. Asterisks denote variables where missing values were mean imputed (mean calculated on training folds only). For NEWS2 missing vital sign values did not contribute to the composite score (score equal to 0), which is a source of uncertainty in our analysis.

| Variable | AUROC (95 % CI) |
| --- | --- |
| Age band | 0.690 (0.642-0.737) |
| Respiration rate* | 0.662 (0.599-0.729) |
| Systolic blood pressure* | 0.586 (0.528-0.645) |
| Heart rate* | 0.537 (0.490-0.585) |
| Temperature* | 0.583 (0.530-0.637) |
| Oxygen Saturation (on air) | 0.772 (0.712-0.833) |
| NEWS2 | 0.715 (0.670-0.760) |
| NEWS2 + age | 0.771 (0.718-0.824)) |
| Oxygen saturation + age | 0.820 (0.785-0.854) |

**Figure 2:**
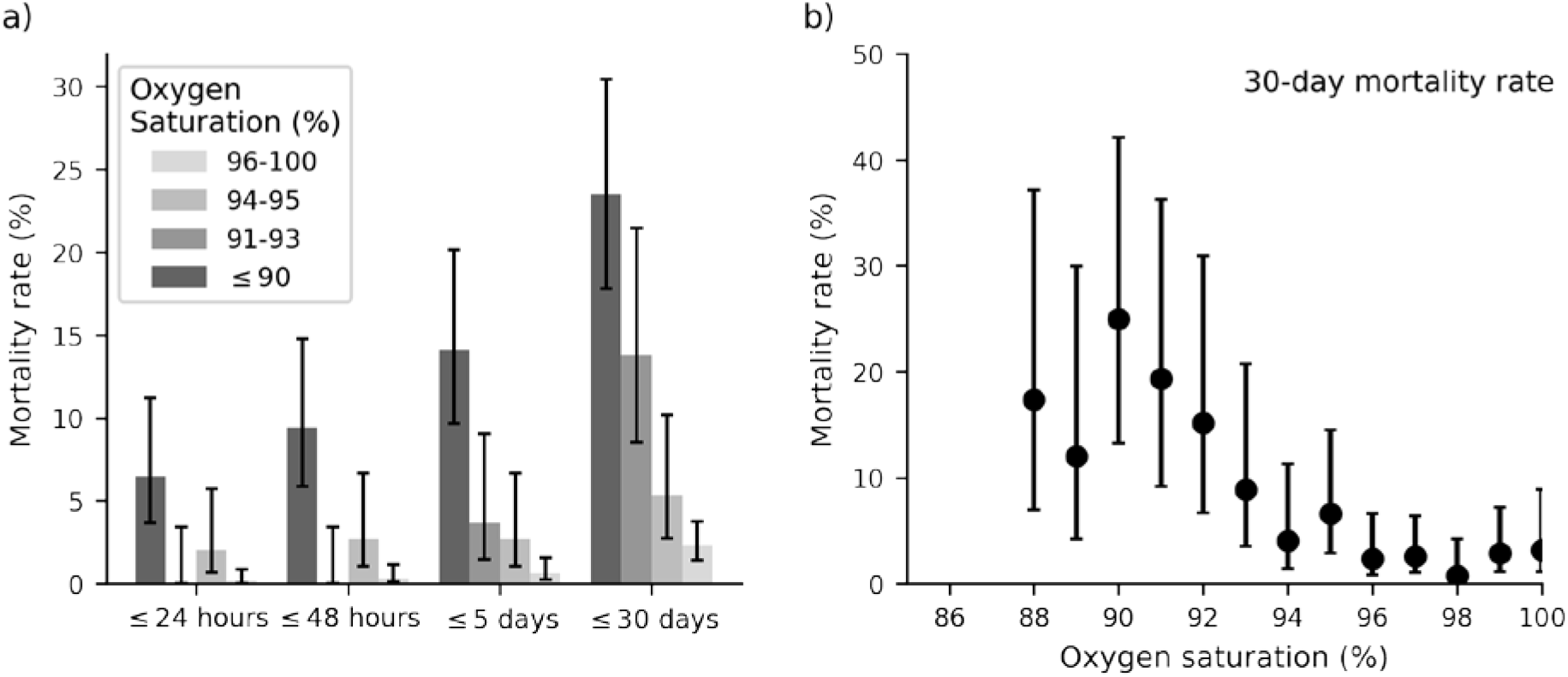
Observed mortality rates as a function of initial oxygen saturation (on air) measurement made by ambulance crews in the community. a) Cumulative mortality rate for aggregated oxygen saturation values (colour, inset legend) for death within 24 hours, 48 hours, 5 days, and 30 days. b) Observed 30-day mortality rate as a function of initial oxygen saturation, highlighting the low-level mortality trend. Points are only shown if there were 10 or more measured oxygen saturation readings at the given level (true for all readings of 88% or greater). Error bars are 95 % confidence intervals estimated using the Wilson score interval.

## DISCUSSION

Community assessment of patients with COVID-19 symptoms using a single initial oxygen saturation on air measurement correlates with both short term (24 hr) and longer term (30 day) mortality rates. Qualitatively, the observed 30-day mortality rate is approximately constant between oxygen saturations of 100 - 96 % (Figure 2 b) and then linearly increases with decreasing oxygen saturation from 95 % to 90 %. Below 90 %, the mortality risk remains high. Although the therapeutic target range for oxygen saturations in the UK is 94-98%,[10] and in the USA is 92-96%,[11] this study suggests that patients at the lower end of this range are still at risk of deterioration in the context of COVID-like symptoms. In Figure 2 a, we demonstrated that even patients with presenting oxygen saturations of 94-95 %, values regarded as within this normal range, had a significantly (p=0.045) higher 30-day mortality rate (5.3 %) than those presenting with oxygen saturations higher than 95 % (30-day mortality rate 2.3 %). Outside this ‘normal’ range, even relatively small decreases in oxygen saturation are markers of increased risk of death and suggest that a lower threshold for hospital conveyance may be necessary for patients who traditionally would be considered to have only minor physiological derangement and otherwise have been left at home.

The sensitivity of home oxygen saturation measurements reflects the percentage of people correctly identified with adverse outcomes. The sensitivity of this parameter for adverse outcome (30-day mortality / high care / ICU admission) decreased as oxygen saturation fell (Table 2). An oxygen saturation ≤ 90% was associated with a relatively low sensitivity of < 0.5. Specificity of identifying an adverse outcome, an indirect measure of unnecessary conveyance to hospital (but also including patients who survived and did not need ICU admissions), increased as oxygen saturations fell. However, it is important to ensure that patients at risk of deterioration are not missed and a degree of over-triage would be necessary to ensure that this was not the case. However, even oxygen saturations at the lower end of the normal range are associated with a risk of deterioration (sensitivity of 94% saturations = 0.713) and it therefore appears that oxygen saturation alone has significant limitations when it is within a normal range. Our data (Table 3) suggests that the addition of age to oxygen saturation measurements may further increase sensitivity to identify risk of mortality at 30 days, but a larger study is required to ascertain whether this is the case.

We also examined co-morbidities known to be risk factors for adverse outcomes with COVID-19 infections.[12] Age was strongly associated with mortality at 30-days, which rose rapidly in those over 60 years of age. COPD patients had almost double the observed 30-day mortality rate compared to those without COPD, but this did not reach statistical significance, likely because of sample size of our study. Contrary to most other studies, diabetes was not a risk factor for adverse outcome in our study, the reason for this is not clear. In total, 21.7% of our sample was diabetic which is similar to that reported in a large UK study of hospitalised COVID-19 patients.[12] Being under cancer care was also not a significant factor for adverse outcome.

Although oxygen saturations as a risk factor for COVID-19 patients on presentation to the Emergency Department are widely reported,[13, 14] [15] the ability of oxygen saturations measured at home to indicate disease severity and the need for hospital conveyance has not been widely reported, presumably because of the challenges in equipping patients with pulse oximeters prior to the onset of any illness. Several studies have used oxygen levels in patients presenting in the Emergency Department as an indicator of the need for hospital admission and others have used the opportunity to send ED patients not requiring admission home with a pulse oximeter for self-monitoring. Oxygen saturations on presentation to the Emergency Department have also been shown to be strongly associated with outcome. The strongest critical illness risk has been shown to be admission oxygen saturation < 88% (OR 6.99).[14] Other studies have shown that even a relatively mildly deranged oxygen saturation of <92% is strongly associated with an increased risk of in-hospital mortality.[16] Conversely, an Emergency Department resting SpO_2_ ≥ 92% as part of discharge criteria can achieve hospital readmission rates as low as 4.6%, [15] suggesting that it may be a safe threshold for discharge in symptomatic patients with mild disease.

Home oxygen saturation monitoring has been used for patients discharged from hospital, both from the ED because their disease was not severe, or from intensive care for convalescence. A small study of patients with COVID-19 discharged from the Emergency Department, reported similar results to ours using subsequent home oxygen saturation monitoring. In these patients, resting home SpO_2_ < 92% was associated with an increased likelihood of re-hospitalization compared to SpO_2_ ≥ 92% (relative risk = 7.0, 95% CI 3.4 to 14.5, p < 0.0001). Home SpO_2_ < 92% was also associated with increased risk of intensive care unit admission.[7]

Oxygen saturation is an integral variable in most critical illness tools that have been used to identify COVID-19 patients requiring hospital admission.[17] NHS England has encouraged the use of the NEWS2 scoring system to identify patients at risk of deterioration. This uses weighted physiological variables of heart rate, systolic blood pressure, oxygen saturation (on air), respiratory rate, temperature and level of consciousness to produce a score that is correlated with risk of deterioration, not only as a general illness score, but specifically in patients with known COVID-19.[18] We therefore compared the ability of isolated oxygen saturations with NEWS2 in our cohort to identify patients at risk of death at 30 days. Using ROC analysis, the AUROC for oxygen saturations alone was 0.772 (95% CI 0.712-0.833) and for NEWS2 was 0.715 (95% CI 0.670-0.760). The addition of age to either oxygen saturation or NEWS2 showed a trend towards improved prediction, but not one that reached statistical significance. These results are consistent with a previous study using NEWS2 scores on hospital admission which has shown an AUROC of 0.822 (95% CI 0.690-0.953) to predict risk of severe disease.[18] The AUROC of NEWS2 in this study may be limited by some missing physiological variables (Table 1). Interestingly, a recent review of 22 prognostic models showed that oxygen saturation on room air and patient age were strong predictors of deterioration and mortality among hospitalised adults with COVID-19 respectively, but no other variables added incremental value to these predictors.[17] We have shown the same for oxygen saturation as a univariate predictor in the pre-hospital setting, and that predictive value does not increase by the addition of other physiological variables.

A number of remote home monitoring models for patients with suspected COVID-19 have been proposed, all of which aim to achieve early identification of deterioration for patients self-managing COVID-19 symptoms at home.[19] It would be expected that the utility of home monitoring would be improved by the ability to measure oxygen saturations, although not all models currently integrate this into their protocols. Our results show that resting oxygen saturations measured in patients with suspected COVID-19 perform on a par with the same measurements taken in the Emergency Department. They therefore suggest that the predictive value of oxygen saturations may be able to be effectively moved to an earlier stage in the disease process and measured while the patient is still at home. Although initial home SpO_2_ may provide a useful marker of disease severity and the need for hospital conveyance, it is clear that it has limited sensitivity and may need to be interpreted as part of an overall assessment of the patient. Some authors have argued that pulse oximetry identified the need for hospitalisation when using a cut-off of 92%,[7] but based on our data (Table 2), approximately one-third of patients with an adverse outcome would be missed using this threshold. We have demonstrated that even patients presenting with oxygen saturations of 94-95 %, which are values regarded as within a normal range, have a higher mortality than those with oxygen saturations higher than 95 %. Even when measured in the Emergency Department, baseline median SpO_2_ was as high as 95.0 % in those with an adverse outcome, compared to 97.0% in those without.[20] It is clear that the relatively low sensitivity of oxygen saturation in those with mildly deranged values limits the utility of this parameter alone in assessing risk of adverse outcome.

This is a relatively small retrospective cohort study with concomitant limitations of sample size. The subjective nature of paramedic classification of symptoms consistent with COVID-19 may have introduced some degree of bias into patients included in the study, as may have the presence of known co-morbidities. Seventeen patients did not have initial oxygen saturations recorded on air (but did have oxygen saturations recorded on oxygen) and were excluded from the data analysis. If this was because they were so obviously hypoxic clinically that ambulance crews immediately administered oxygen without an initial reading on air (or were constantly on home oxygen treatment), the ability of oxygen saturations to indicate risk of deterioration is likely to have been underestimated in this study. Patients on palliative care pathways were also removed from the study cohort but are obviously likely to be more susceptible to deterioration from COVID, irrespective of any alternative care pathway.

With waves of COVID-19 regularly overwhelming ambulance services and hospital services, there is an urgent need to optimise the identification of patients at risk of deterioration. This will enable more patients to safely be managed in the community and only referred to hospital once their clinical symptoms and physiological signs suggest a risk of deterioration and the need for hospital care. This is particularly needed for the majority of patients who have mild to moderate symptoms where it is not clear where community or hospital management is most appropriate. Home pulse oximetry is becoming relatively cheap and easily accessible for the public and may be a relatively cost-effective tool in the safe community management of these patients, perhaps focussed on those with significant co-morbidities who are at higher risk. The utility of remote monitoring systems (or the COVID virtual ward) has been an increasingly studied subject, and there is growing evidence that remote monitoring can facilitate more streamlined approaches to the delivery of patient care, especially in pulmonary disease.[6] Further prospective studies are required to understand the utility of home pulse oximetry, but this study suggests that it may have the potential to significantly contribute to the safe and appropriate management of these patients in the community with timely referral to hospital when indicated.

## Conclusions

We have demonstrated that even relatively minor derangements in peripheral oxygen saturation are an early warning of potential deterioration and oxygen saturation would appear to have potential to be a key physiological variable that together with other clinical signs may be able to identify patients at risk of deterioration.

## Data Availability

Due to information governance concerns, the data will not be made public. However, it will be made accessible via reasonable request to the corresponding author.

## Acknowledgements

We thank Simon Mortimore and Philip King from South Central Ambulance Service and Zoe Cameron from Hampshire Hospitals NHS Foundation Trust for their assistance in data extraction and analysis.

This report includes independent research funded by the National Institute for Health Research Applied Research Collaboration Wessex. The views expressed in this publication are those of the author(s) and not necessarily those of the National Institute for Health Research or the Department of Health and Social Care.

## Governance and ethics approval

Regulatory and ethical approval for the study were provided by the Health Research Authority (REC reference 20/HRA/5445) and by the University of Southampton Ethics Committee (REF ERGO/61242). NHS England and NHS Improvement have been given legal notice by the Secretary of State for Health and Social Care to support the processing and sharing of information to help the COVID-19 response under Health Service Control of Patient Information Regulations 2002 (COPI). This is to ensure that confidential patient information can be used and shared appropriately and lawfully for purposes related to the COVID-19 response. Data were extracted from medical records by clinicians providing care for the patients and an anonymised extract of the data were provided to the team at the University of Southampton.

## Supplementary Information

**Supplementary Table 1:** Example of predictor which combines oxygen saturation (columns) and age group (rows). Each element of the table displays the observed 30-day mortality rate (%) for the respective group. Practically, this is used as a look-up table to predict a patient’s 30-day mortality risk. For example, a patient of 75 years old who had an oxygen saturation of 88 % (measured by a paramedic in the community after a 999 or 111 call) would be assigned a 30-day mortality risk of 24.6 %. This heuristic model is only a demonstration to show the potentially increased predictive power of combining oxygen saturation with age. It is important to note this table is only an example and the numbers do not reflect that used in the main manuscript (Table 3) – they are the mean of ten predictors made from table 3 with mortality rates calculated for 10 different views of the dataset seen in the cross validation process.

|  |  | Oxygen Saturation (%) |  |  |  |
| --- | --- | --- | --- | --- | --- |
|  |  | ≤90 | 90-93 | 94-95 | 96-100 |
| Age group (years) | 18-59 | 5.4 | 4.3 | 0.0 | 0.0 |
|  | 69-79 | 24.6 | 8.1 | 2.0 | 1.9 |
|  | 80+ | 32.8 | 22.4 | 10.4 | 5.2 |

**Supplementary Table 2:** Example of predictor which combines NEWS2 (columns) and age group (rows). Each element of the table displays the observed 30-day mortality rate (%) for the respective group. Practically, this is used as a look-up table to predict a patients 30-day mortality risk in the main text. For example, a patient of 75 years old who had an NEWS2 if 6 (measured by a paramedic in the community after a 999 or 111 call) would be assigned a 30-day mortality risk of 15.9 %. This heuristic model is only a demonstration to show the potentially increased predictive power of combining NEWS2 with age. It is important to note this table is only an example and the numbers do not reflect that used in the main manuscript (Table 3) – they are the mean of ten predictors made from this table with mortality rates calculated for 10 different views of the dataset seen in the cross-validation process

|  |  | NEWS2 score |  |  |  |
| --- | --- | --- | --- | --- | --- |
|  |  | 0-2 | 3-5 | 6-8 | 9+ |
| Age band (years) | 18-59 | 0 | 1.0 | 1.5 | 2.4 |
|  | 69-79 | 1.3 | 5.2 | 15.9 | 14.3 |
|  | 80+ | 3.7 | 13.6 | 23.5 | 25.5 |

